# Gender Moderates the Mediating Pathway between Social Deprivation, Body Mass Index and Physical Activity

**DOI:** 10.1101/2022.04.18.22273963

**Authors:** Silvio Maltagliati, Ilyes Saoudi, Philippe Sarrazin, Stéphane Cullati, Stefan Sieber, Aïna Chalabaev, Boris Cheval

## Abstract

Physical activity is unequally practiced across populations: relative to more privileged individuals, deprived people are less likely to be physically active. However, pathways underlying the association between deprivation and physical activity remain overlooked. Here, we examined whether the association between several indicators of deprivation (administrative area deprivation and self-reported individual material and social deprivation) and physical activity was mediated by body mass index (BMI). In addition, consistent with an intersectional perspective, we tested whether this mediating pathway was moderated by participants’ gender and we hypothesized that the mediating effect of BMI would be stronger among women, relative to men. We used two independent large-scale studies to test the proposed pathways cross-sectionally and prospectively. In a first sample composed of 5,723 British adults (Study 1), BMI partly mediated the cross-sectional association between administrative area deprivation and self-reported physical activity. Moreover, relative to men, the detrimental effect of deprivation on BMI was exacerbated among women, with BMI mediating 3.1% of the association between deprivation and physical activity among women (vs 1.5% among men). In a second sample composed of 8,358 European older adults (Study 2), our results confirmed the findings observed in Study 1: BMI partly mediated the prospective association between perceived material and social deprivation and self-reported physical activity. Moreover, compared to men, the effect of deprivation on BMI was more pronounced among women, with BMI respectively mediating 8.1% and 3.4% of the association between material and social deprivation and physical activity among women (vs 1.3% and 1.2% among men). These findings suggest that BMI partly explained the detrimental association between deprivation and physical activity, with this effect being stronger among women. Our study highlights the need to further consider how gender may shape the mechanisms behind the association between disadvantaged socio-economic circumstances and physical activity.

---

In most developed countries, a significant decrease in mortality has been observed across the twentieth century (Oeppen & Vaupel, 2002). Yet, this trend is not equally distributed across the population (Lallo & Raitano, 2018; Remund et al., 2019) – the health gap between advantaged and disadvantaged individuals has, at best, maintained over time, or rather widened for some outcomes (Dowd et al., 2011; Tawiah et al., 2022). Disparities in the adoption of health behaviors among middle-aged and older adults, such as physical activity, may partially explain why social health inequalities are broadening (Petrovic et al., 2018; Stringhini, 2010). While the benefits of being physically active are extensive (Rhodes et al., 2017), disadvantaged populations consistently report lower levels of physical activity during leisure time (O’Donoghue et al., 2020). In order to reduce health inequalities, understanding the mechanisms underlying the association between disadvantaged socio-economic circumstances and health behaviors is needed.

Amongst the socio-economic circumstances linked with physical inactivity is deprivation, a multidimension construct which is defined as a deficiency in basic material and social needs (Myck et al., 2015). It encapsulates, but does not restrict to, traditional indicators of socio-economic circumstances (e.g., income, education), by focusing on access to material (i.e., access to goods) and social opportunities (i.e., access to social support), over and beyond individuals’ socio-economic position. Compared with more privileged individuals, deprived people have been shown to be less physically active during leisure time (Farrell et al., 2014; Fox et al., 2011). Yet, the pathways through which deprivation relates to lower physical activity remain overlooked (Diez Roux, 2016). Beyond environmental (e.g., access to facilities, walkability or adverse neighborhood conditions, air pollution) (Chaparro et al., 2018; Hillsdon et al., 2007; Schneider et al., 2019), cultural (e.g., class habitus) (Bourdieu, 1979; Cockerham et al., 2020; Pampel et al., 2010), or cognitive factors (e.g., motivations, cognitive functions) (Cheval et al., 2019; Hagger & Hamilton, 2020), body mass index (BMI) stands as one potential mediating candidate, though empirical evidence remains lacking.

Indeed, on the one hand, deprivation has been found to be associated with a higher BMI (Bann et al., 2017; Matheson et al., 2008), potentially because deprived obesogenic environments encourage higher food intake and restrict the access to affordable and nutritious food (Atanasova et al., 2022). Moreover, at the individual level, perceiving deprivation can tax the cognitive ‘bandwidth’, which leads to a weaker ability to self-regulate one’s own behavior and to resist giving into unhealthy food consumption (Beenackers et al., 2016; Mesler et al., 2022). On the other hand, yet bi-directional associations between physical activity and BMI are proposed (see the Discussion section), a higher BMI has been repeatedly associated with a lower participation in physical activity (e.g., Cassidy et al., 2017; Ekelund et al., 2017). In sum, while deprivation has been associated with a higher BMI, which has, in turn, been shown to restrain engagement in PA, whether BMI mediated the association between deprivation and physical activity has never been formally tested – a first knowledge gap that this study aims to fill.

Beyond the effect of deprivation on physical activity through BMI, an intersectional perspective further suggests that socio-economic factors may combine, and even widen inequalities in health behaviors (Bauer, 2014; Bowleg, 2012). For example, socioeconomic status and gender were found to interact when predicting health condition and physical activity, with the detrimental effects of a disadvantaged socio-economic background on health condition (Mackenbach et al., 1999) and on physical activity (Chalabaev et al., 2022; Cheval et al., 2018) being especially pronounced among women. Here, we postulate that gender could moderate the mediating pathway between deprivation, BMI and physical activity in two distinct ways. First, gender may moderate the association between deprivation and BMI (i.e., first component of the mediating pathway) as, relative to men, the association between deprivation and BMI has been shown to be exacerbated among women (Bann et al., 2017; Matheson et al., 2008). Second, gender may also moderate the relationship between BMI and physical activity (i.e., second component of the mediating pathway): a higher BMI may represent a stronger barrier to physical activity among women, relative to men. Particularly, women with a higher BMI are more exposed to stigma than their male counterparts (Judge & Cable, 2011), which may in turn restrain their engagement in physical activity (Rojas-Sánchez et al., 2021; Vartanian & Shaprow, 2008). However, at the time of writing, no study has investigated whether and how gender moderates the mediation between deprivation, BMI and physical activity.

The aim of the current study was twofold. First, we aimed to investigate whether BMI mediates the association between deprivation and physical activity. Second, we aimed to test whether gender moderates this mediating pathway. We hypothesized that BMI would mediate the association between deprivation and physical activity (H1). We further hypothesized that gender would moderate this mediating pathway – the mediating role of BMI on the relationship between deprivation and physical activity would be stronger among women than among men (H2). We thus examined whether gender moderated the association between deprivation and BMI (i.e., first component of the mediating pathway) and/or the association of BMI with physical activity (i.e., second component of the mediating pathway). To test these hypotheses, we relied on two independent large-scale samples in which different indicators of deprivation were assessed. Beyond assessing the robustness of results in two independent samples, it allowed us to examine whether associations between deprivation and physical activity differed across different facets of socio-economic circumstances (Chalabaev et al., 2022). In Study 1, deprivation was conceptualized at the “administrative area” level, meaning that this indicator was built upon participants’ area of residence and aggregated on the basis of several indicators related to the area (Ellaway et al., 2012). In Study 2, deprivation was assessed at an “individual” level, using two self-reported perceived indicators: material deprivation (e.g., having access to necessary groceries, clothes) and social deprivation (e.g., relatedness with people in the local area, vandalism) (Myck et al., 2020).

## Study 1

This first study aimed to provide cross-sectional evidence on the mediating role of BMI on the association between area deprivation and physical activity. Moreover, it examined whether gender moderates this mediating pathway.

### Methods

#### Participants and procedure

Data were retrieved from a publicly available dataset (https://osf.io/uzbw9/). The study was extensively described elsewhere (Steele et al., 2021). Briefly, British adults participating in different exercise referral schemes across the country were invited to complete a single-time questionnaire assessing sociodemographic variables, as well as variables related to physical activity and health indicators. Data were screened, processed and cleaned by the original research team (Steele et al., 2021). We included individuals having complete measures on the variables of interest (i.e., area deprivation, BMI, physical activity and gender). We also excluded participants who answered “transgender” or “unknown” on the gender item (n = 32), resulting in a total of 5,723 participants (66% women, mean age = 53 ± 15 years).

#### Measures

##### Predictor

*Area deprivation* was assessed using the multiple index of deprivation. Participants self-reported their postcode, which was used to determine the multiple index of deprivation for each participant, depending on their living area. This measure aggregates seven weighted domains of deprivation (i.e., income, employment, health deprivation and disability, education, crimes, barriers to housing and services, living environment), which higher values reflecting a higher level of deprivation. This scoring procedure allows to rank local areas of the United Kingdom relative to their level of deprivation (32,844 areas in total, with an average population of ∼1500 individuals), as well as to predict health-related outcomes in previous research (Farrell et al., 2014).

##### Mediating variable

*BMI* (in kg/m^2^) was calculated based on participants’ self-reported height (in cm) and weight (in kg).

##### Outcome

*Physical activity* was measured using the short form of the International Physical Activity Questionnaire (IPAQ) (Craig et al., 2003). In reference to the last seven days, participants were asked to report the number of days during which they practiced vigorous and moderate physical activity and walking (for at least 10 minutes). Then, they indicated how much time they usually spent doing each of these activities (in hours and minutes). These scores were used to estimate total physical activity in Metabolic Equivalent of Task/minute (Craig et al., 2003).

##### Moderating variable

*Gender* was self-reported by participants, with retained answering options being: “female” and “male”.

*Confounders* were age (in years), employment status (categorical variable) and ethnicity (categorical variable) (see Steele et al., 2021). Based on available data in the sample, these confounders were selected after we had specified our conceptual model in which we aimed to identify variables which could be related to the predictor, the mediator and the outcome (Lipsky & Greenland, 2022).

#### Statistical analyses

All analyses were computed on R ®, version 4.0.4. Continuous variables (i.e., area deprivation, BMI, physical activity) were centered and scaled, while gender was contrast-coded (−0.5 for men and 0.5 for women). Simple mediation models were first computed by entering area deprivation, BMI and physical activity as the predictor, the mediating variable and the outcome, respectively. Then, a moderated mediation analysis was specified by adding gender as the moderating variable on this mediating pathway.

To test the mediating pathway between area deprivation, BMI and physical activity, we adopted two complementary approaches (Yzerbyt et al., 2018). First, as proposed by the index approach, mediation analyses were conducted based on the distribution-of-the product framework in order to estimate confidence intervals around the indirect effects (Hayes, 2015). A Monte Carlo 95% confidence interval (95% CI) around the indirect effect index which did not contain 0 is assumed to indicate a significant indirect effect (with 5,000 simulations). Second, as proposed by the component approach, to confirm the significance of the indirect effects, the two paths of the indirect effect were tested (i.e., from the predictors to the mediating variable [*a* path] and from the mediating variable to the dependent variable [*b* path]), with an indirect effect being supported when both paths are significant (*p* < .05) (Yzerbyt et al., 2018).

To examine the moderating effect of gender on this mediating pathway, we also relied on these two approaches. Specifically, as proposed by the index approach, we first tested whether the Monte Carlo 95% CI contained 0 for the index of the moderated indirect effect, with an interval not containing 0 being indicative of a significant moderated mediation. Then, as suggested by the component approach, we examined whether gender moderated the association between area deprivation and BMI (*a* path) or the association between BMI and physical activity (*b* path) (Muller et al., 2005). Simple slopes analyses were further computed to decompose potential moderating effects.

Models were specified using the package mediation (Tingley et al., 2014) and were inspected using the package performance (i.e., linearity and normality of residuals, homogeneity of variance, undue influence).

### Results

Descriptive statistics and univariate associations are provided in Table 1 and full results are provided in Supplementary Material (Table S1).

**Table 1.** Descriptive statistics and univariate associations.

| **Study 1** |  |  |  |
| --- | --- | --- | --- |
| | Mean (SD) | Range | $\chi^2/r, p$-value |
| **Physical activity** (in MET/min) | 1174 (2066) | 0 – 17118 | - |
| **Area deprivation** | 18.46 (13.66) | 0.90 – 70.89 | $r = -.34, p < .001$ |
| **BMI** | 30.81 (6.80) | 12.8 – 69.4 | $r = -.12, p < .001$ |
| **Gender** (N, % of women) | 3782, 66% | - | $r = .14, p < .001$ |
| **Study 2** |  |  |  |
| **Physical activity** |  |  |  |
| Physically active (N, %) | 6041 (72%) | - | - |
| Physically inactive (N, %) | 2316 (28%) | - | - |
| **Material deprivation** (from 0 to 1) | 0.10 (0.16) | 0 – 1 | $r = -.15, p < .001$ |
| **Social deprivation** (from 0 to 1) | 0.16 (0.13) | 0 – 0.83 | $r = -.23, p < .001$ |
| **BMI** | 26.98 (4.64) | 12.8 – 74.7 | $r = -.08, p < .001$ |
| **Gender** (N, % of women) | 4662 (56%) | - | $\chi^2 = 19.22, p < .001$ |

*Note.* SD: standard deviation. Univariate associations with physical activity were computed using correlations ( $r$ ) and chi-square tests ( $\chi^2$ ) for continuous and categorical variables, respectively. For gender, being a man (vs a woman) was positively associated with physical activity level (Study 1) or odds of being classified as physically active (vs inactive) (Study 2).

Results of the simple mediation model showed a significant indirect effect of area deprivation on physical activity through BMI (index of the indirect effect = -0.0091, 95% CI = [-0.0150; -0.0044], *p* < .001). Specifically, area deprivation was positively associated with BMI (Model 1: b = .19, 95% CI = [.16; .22], *p* < .001) and, in turn, higher BMI was associated with lower physical activity (Model 2: b = -.05, 95% CI = [-.07 -.03], *p* < .001). The direct effect of area deprivation on physical activity remained significant after adjustment for BMI (Model 2: b = -.29, 95% CI = [-.32; -.26], *p* < .001).

Results of the moderated mediation analyses showed that the indirect effect of area deprivation on physical activity through BMI was moderated by gender (index of the moderated indirect effect = -.0071, 95% CI = [-.0095; -.0047], *p* < .001) (Figure 1). Relative to men (index of the indirect effect = -.0040, 95% CI = [-.010; -.001], *p* = .056), the indirect effect was more than twofold stronger among women (index of the indirect effect = -.0113, 95% CI = [-.0180; - .0048], *p* < .001). This stronger indirect effect was accounted by the significant moderating effect of gender on the association between area deprivation and BMI (*a* path) (Model 1: b = .17, 95% CI = [.13; .21], *p* < .001). Simple slope analysis revealed that the association of area deprivation with BMI was stronger among women (b = .24, 95% CI = [.20; .28], *p* < .001), relative to men (b = .07, 95% CI = [.03; .11], *p* < .001) (Figure 2). However, gender did not significantly moderate the strength of the association between BMI and physical activity (*b* path) (*p* = 0.505). Among women, BMI mediated around 3.1% of the association between area deprivation and physical activity, against 1.5% among men.

**Figure 1.**
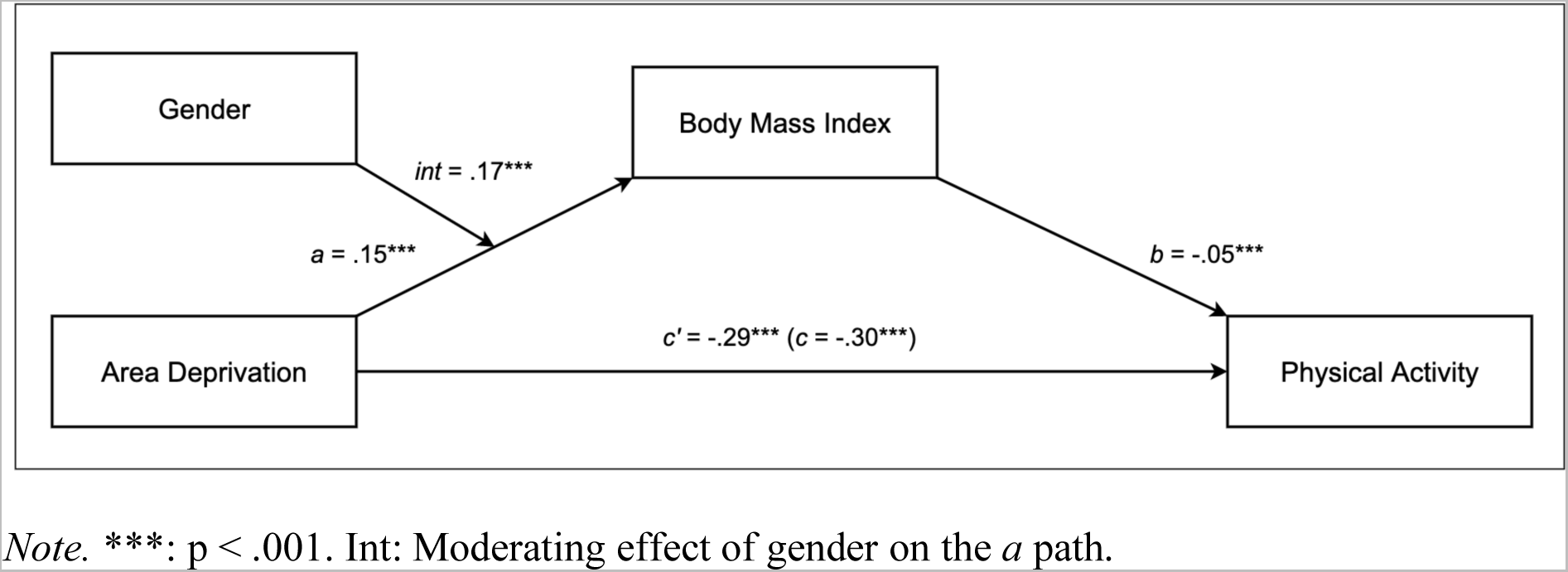
Moderated mediation model for Study 1.

**Figure 2.**
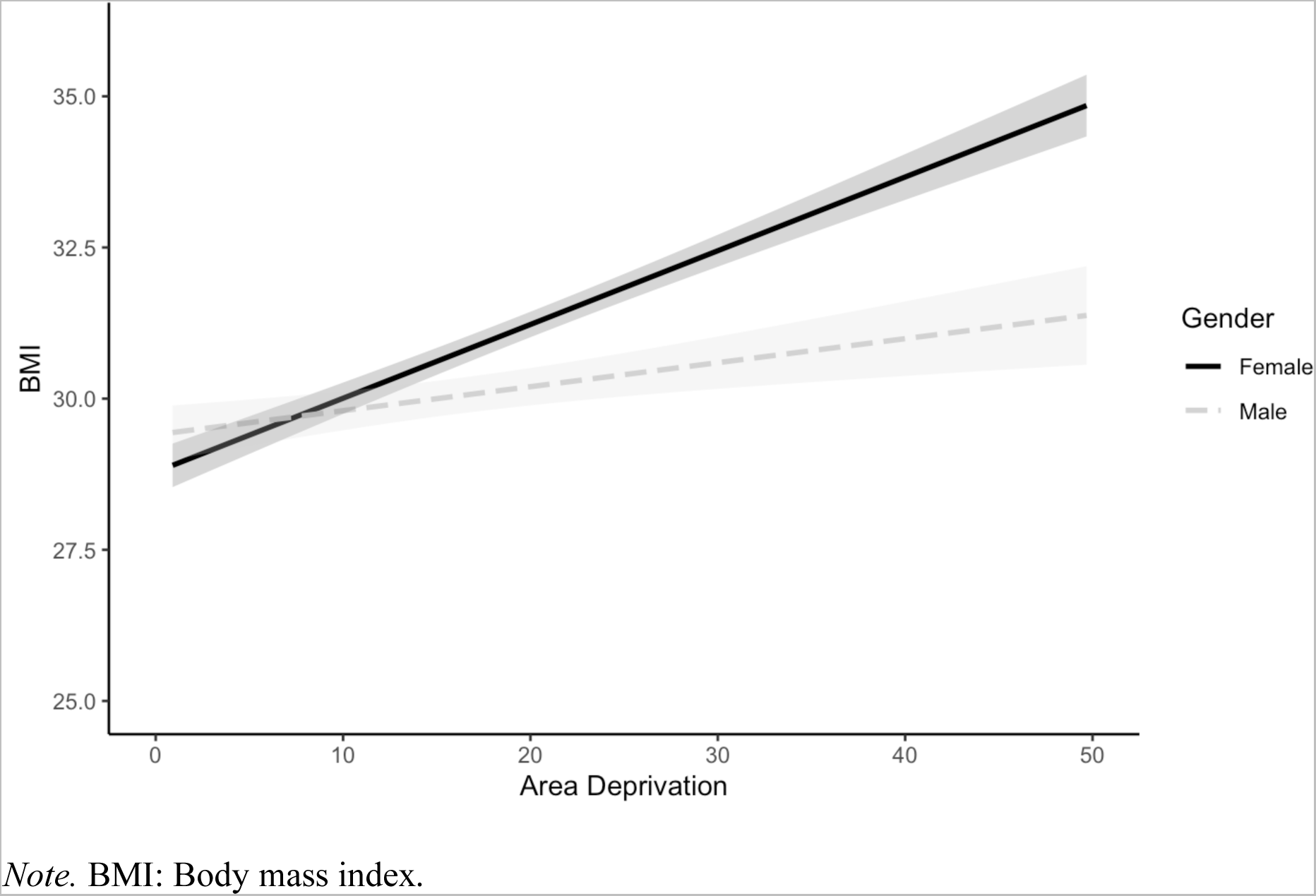
Simple slopes for the association between area deprivation and BMI, depending on gender (Study 1).

### Discussion

These findings suggested that, beyond individual factors (i.e., age, ethnicity and employment status), BMI partly mediated the cross-sectional association between area deprivation and physical activity. Of note, this mediating pattern depended on gender: relative to men, the detrimental association of area deprivation with BMI was exacerbated among women (i.e., first component of the mediating pathway). However, at least three features limited these conclusions that could be drawn from this study. First, area deprivation, BMI and physical activity were measured on a single timepoint, which stood in contrast with the principle of temporal precedence in mediation analyses (Yzerbyt et al., 2018). Second, the composite nature of the multiple index of deprivation prevented us to disentangle the different facets of deprivation (e.g., social versus material deprivation). Yet, a recent study has underlined potential distinct associations of social and material deprivation with physical activity (Chalabaev et al., 2022). Moreover, this index only reflected deprivation at a crude area level. Notably, some non-deprived individuals may live in deprived areas, and many deprived people may live in non-deprived areas (e.g., Farrell et al., 2014), which increases potential misclassification bias. Moreover, given that “perceived” deprivation constitutes a proximal driver of health behaviors (Singh-Manoux et al., 2005), examining the effects of self-reported deprivation on physical activity would provide a complementary approach to Study 1. Finally, due to missing data, we were unable to adjust our models to additional confounding variables (e.g., education or income). Replicating our findings with a time-lagged assessment of variables, material and social measures of deprivation at the individual-level and the adjustment for additional confounders was thus warranted.

## Study 2

This second study aimed to address the abovementioned limitations by examining whether BMI mediated the association between deprivation with physical activity using 1) a prospective design (i.e., physical activity was measured about two and four years after the mediator and the predictor, respectively), 2) complementary indicators of material and social deprivation at the individual-level and 3) adjusting for additional variables that may confound the associations in the hypothesized model. Moreover, while the first sample was only composed of middle-aged British adults involved in exercise referral schemes, the sample of this Study 2 relies upon a more representative sample of older European adults.

### Methods

#### Participants and procedure

Data were drawn from the Survey of Health, Ageing and Retirement in Europe (SHARE), a longitudinal population-based study on European adults 50 years of age or older. All procedures and measures of SHARE were described in detail elsewhere (Börsch-Supan et al., 2013). In short, data were collected every two years on eight waves, between 2004 and 2019 using computer-assisted personal interviewing (CAPI) in participants’ homes. On wave 5 (in 2013), material and social deprivation were assessed using a devoted set of items. Two and four years later respectively, in wave 6 and in wave 7, BMI and physical activity were measured. We focused on these three waves of measurement as it provides a short-time prospective window to estimate the mediating pattern between deprivation, BMI and physical activity. To be included in the study, participants had to be between 50 years and 96 years old and had completed measures of deprivation on wave 5, of BMI on wave 6 and of physical activity on wave 7.

In total, 8,358 individuals were included in the study (56% of women, mean age = 70 ± 8 years). In this sample, 645 participants (8% of the full sample) were classified as severely deprived (i.e., allocated above the 75th percentile of the full sample distribution on both social and material deprivation), and 6,041 individuals (72% of the full sample) were categorized as physically active.

#### Measures

##### Predictors

*Perceived material and social deprivation* were measured by a questionnaire in wave 5. For material deprivation, participants answered on 11 items focusing on different dimensions of the economic circumstances of households (e.g., affording to buy fruit and meat, necessary groceries, clothes, shoes, to go on holidays, to receive health care). An aggregated weighted indicator of material deprivation was computed by averaging the different items (Bertoni et al., 2015). The score ranged from continuous values between 0 and 1, with higher values reflecting a higher material deprivation.

*For social deprivation*, participants answered on 15 items focusing on different dimensions of everyday life participation (e.g., helpfulness of people in the local area, skills in reading or in technologies), as well as on the perceived quality of the neighborhood (e.g., access to shops, banks and health structures, vandalism). Similar to material deprivation, items were averaged into a single weighted index of social deprivation (Myck et al., 2015). The score ranged from continuous values between 0 and 1, with higher values reflecting a higher social deprivation.

These indicators of perceived deprivation have been used to predict health-related outcomes, including perceived health status, difficulties in mobility, risks of mortality and well-being (Myck et al., 2020; Terraneo, 2021).

##### Mediating variable

*BMI* (in kg/m^2^) was computed based on participants’ self-reported height and weight in wave 6.

###### Outcome

*Physical activity* was self-reported by questionnaire in wave 7, using the two following questions: “How often do you engage in vigorous physical activity, such as sports, heavy housework, or a job that involves physical labor?” and “How often do you engage in activities that require a low or moderate level of energy such as gardening, cleaning the car, or doing a walk?”. Participants answered using a four-point scale with the following options: *Hardly ever, or never*; *One to three times a month*; *Once a week*; *More than once a week*. Participants who did not answer “more than once a week” to either item were classified as physically inactive (coded 0), while other participants were classified as physically active (coded 1). As described in previous research (Cheval et al., 2018, 2019), this strategy reduces the potential misclassification bias which would lead to physically inactive participants being incorrectly classified as physically active.

This two-item scale has been found to be robustly associated with health outcomes, including, for example, chronic diseases (de Souto Barreto et al., 2017), cognitive functions (Cheval et al., 2019, 2020), depressive symptoms (Boisgontier et al., 2020), diabetes (Cheval et al., 2021) or risks of hospitalization following COVID-19 infections (Maltagliati et al., 2021). Moreover, in the context of longitudinal large-scale surveys, similar one-or two-items scales have provided acceptable validity regarding the assessment of physical activity (Milton et al., 2011), including among older adults (Gill et al., 2012)

###### Moderating variable

*Gender* was measured in wave 5, with participants reporting being either “female” or “male”.

*Confounders* included participants’ age, country of residence, education, income and occupation status (e.g., Cheval et al., 2018). Education, income and occupation status were measured when participants were first included in the study. These confounders were selected after we had specified our conceptual model in which we aimed to identify variables which could be related to both the predictor, the mediator and the outcome (Lipsky & Greenland, 2022).

##### Statistical analyses

As in Study 1, simple and moderated mediation analyses were computed, with material deprivation and social deprivation being separately used as predictors (*r* = .37, *p* < .001). We followed the same preliminary steps than previously described in Study 1 (i.e., standardizing and contrast-coding continuous and dichotomous variables, respectively). In Study 2, however, the outcome was dichotomous (i.e., physically inactive vs physically active individuals). Consequently, logistic regression models were conducted to test the association of predicting variables with the outcome. We report unstandardized beta coefficients (*b*), as well as odds-ratios (OR), which were calculated by exponentiating *b*.

### Results

Descriptive statistics and univariate associations are provided in Table 1 and full results are provided in Supplementary Material (Tables S2 and S3).

#### Material deprivation

Results of the simple mediation model showed a significant indirect effect of material deprivation on physical activity through BMI (index of the indirect effect = -.0020, 95% CI = [-.0032; -.0008], *p* = .004). Specifically, material deprivation was positively associated with BMI (Model 1: b = .06, 95% CI = [.02; .09], *p* < .001) and, in turn, higher BMI was associated with lower odds of being physically active (vs inactive) (Model 2: b = -.19, 95% CI = [-.25 - .14], OR = .82, 95% CI = [0.78; 0.87], *p* < .001). The direct effect of material deprivation on physical activity remained significant after adjustment for BMI (Model 2: b = -.16, 95% CI = [-.23; -.08], OR = .85, 95% CI = [0.79; 0.92], *p* < .001).

Results of the moderated mediation analyses showed that the indirect effect of material deprivation on physical activity through BMI was moderated by gender (index of the moderated indirect effect = -.0032, 95% CI = [-.0040; -.0024;], *p* < .001) (Figure 3A). Relative to men (index of the indirect effect = -.0003, 95% CI = [-.0023; .0016], *p* = .780), the indirect effect was significant and stronger among women (index of the indirect effect = -.0030, 95% CI = [- .0050; -.0012], *p* < 001). This stronger indirect effect was accounted by the significant moderating effect of gender on the association between material deprivation and BMI (*a* path) (Model 1: b = .08, 95% CI = [.03; .13], *p* < .001). Simple slope analysis revealed that the association of material deprivation with BMI was stronger among women (b = .09, 95% CI = [.05; .13], *p* < .001), relative to men (b = .01, 95% CI = [-.03; .05], *p* = .750) (Figure 4A).

**Figure 3.**
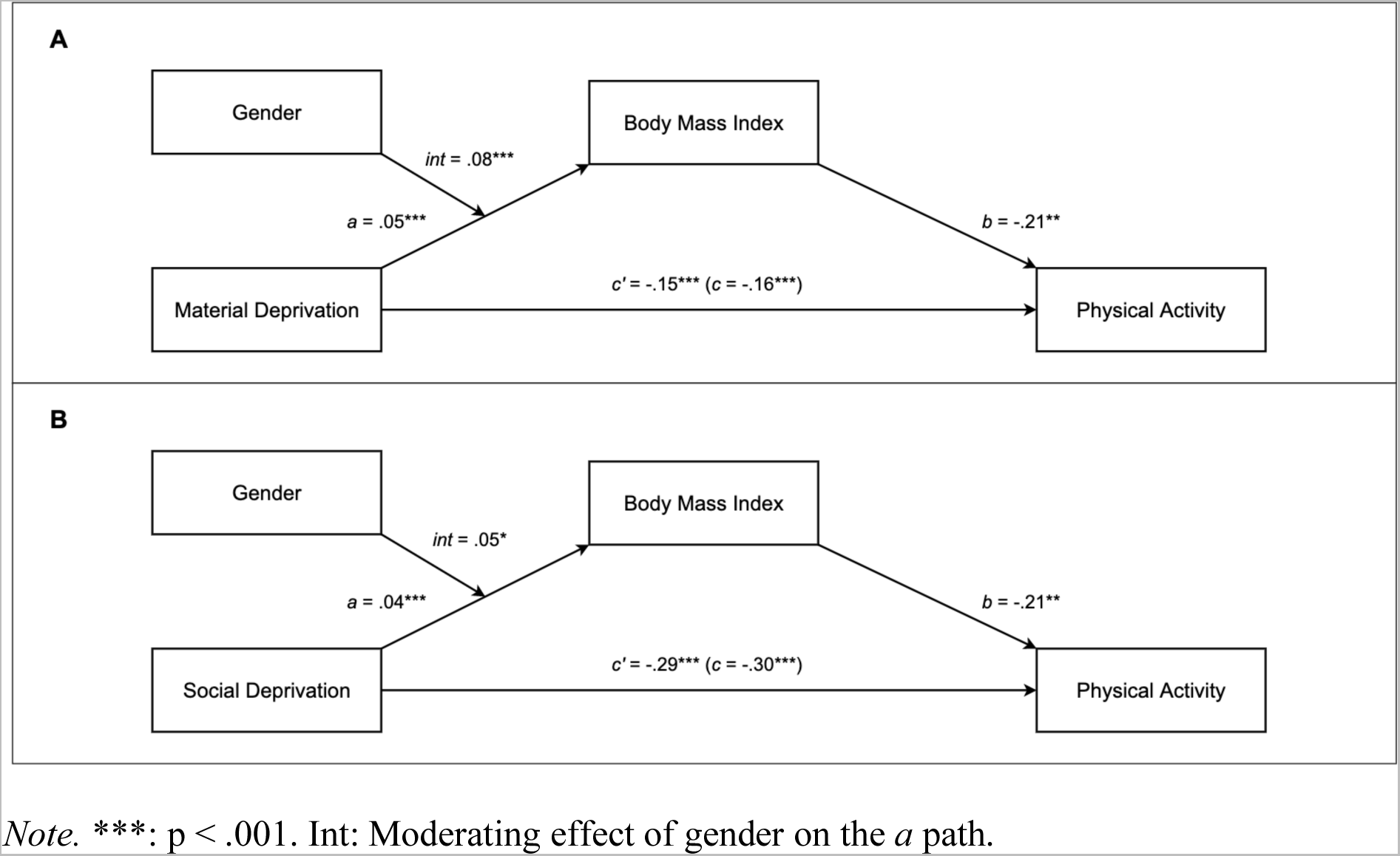
Moderated mediation models with material deprivation (A), social deprivation (B) for Study 2.

**Figure 4.**
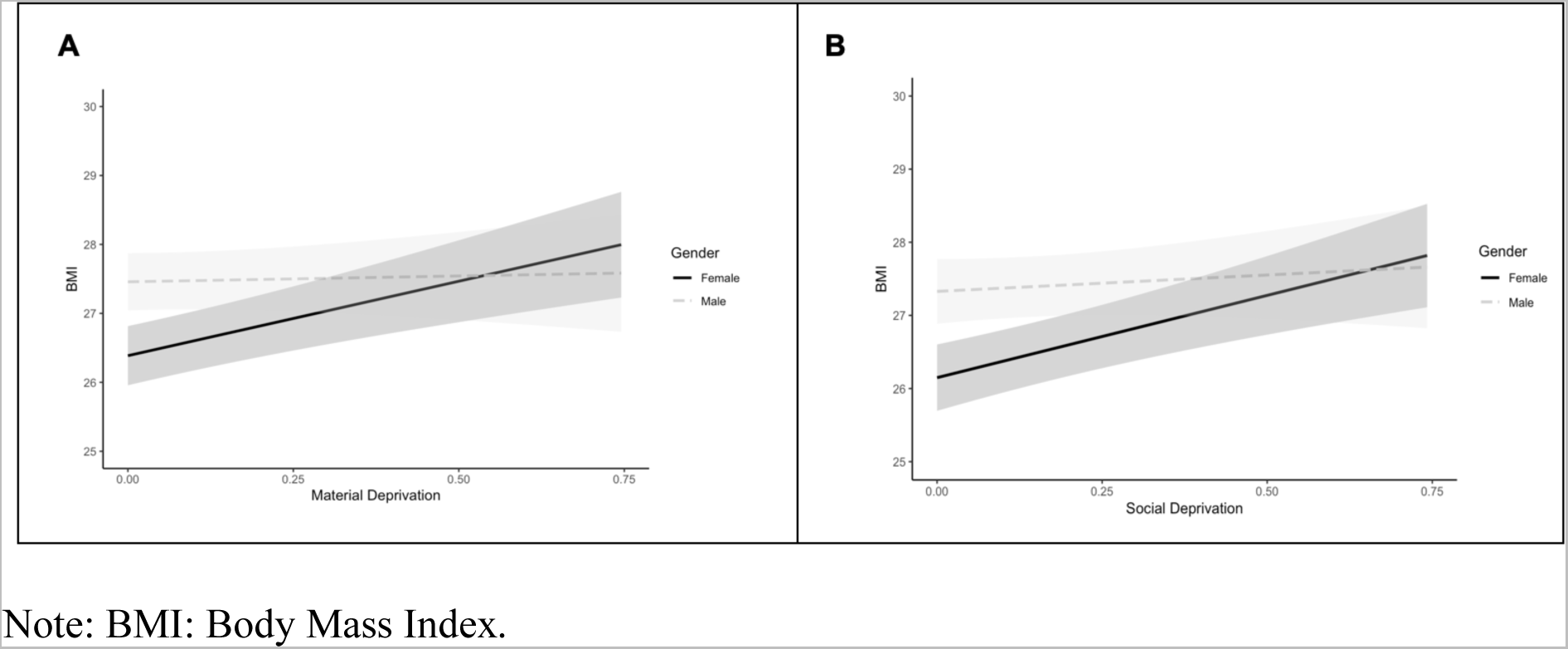
Simples slopes for the interaction of material deprivation (A) and social deprivation with gender on BMI (Study 2).

Moreover, as in Study 1, gender did not significantly moderate the strength of the association between BMI and physical activity (*b* path) (*p* = 0.220). Among women, BMI mediated around 8.1% of the association between material deprivation and physical activity, against 1.3% among men.

#### Social deprivation

Results of the simple mediation model showed a significant indirect effect of social deprivation on physical activity through BMI (index of the indirect effect = -.0015, 95% CI = [-.0026; -.0005], *p* = .002). Specifically, social deprivation was positively associated with BMI (Model 1: b = .04, 95% CI = [.02; .06], *p* < .001) and, in turn, higher BMI was associated with lower odds of being physically active (vs inactive) (Model 2: b = -.19, 95% CI = [-.24 -.14], OR = .83, 95% CI = [0.78; 0.87], *p* < .001). The direct effect of social deprivation on physical activity remained significant after adjustment for BMI (Model 2: b = -.30, 95% CI = [-.36; - .24], OR = .74, 95% CI = [0.70; 0.79], *p* < .001).

Results of the moderated mediation analyses showed that the indirect effect of social deprivation on physical activity through BMI was moderated by gender (index of the moderated indirect effect = -.0045, 95% CI= [-.0058; -.0033], *p* < .001) (Figure 3B). Relative to men (index of the indirect effect = -.0006, 95% CI = [-.0022; .0009], *p* = .520), the indirect effect was significant and stronger among women (index of the indirect effect = -.0022, 95% CI = [-.0038; -.0010], *p* < .001). This stronger indirect effect was accounted by the significant moderating effect of gender on the association between social deprivation and BMI (*a* path) (Model 1: b = .05, 95% CI = [.01; .10], *p* = .015). Simple slope analysis revealed that the association of social deprivation with BMI was stronger among women (b = .07, 95% CI = [.05; .09], *p* < .001), relative to men (b = .01, 95% CI = [-.03; .05], *p* = .460) (Figure 4B). Moreover, as in Study 1, gender did not significantly moderate the strength of the association between BMI and physical activity (*b* path) (*p* = 0.233). Among women, BMI mediated around 3.4% of the association between social deprivation and physical activity, against 1.2% among men.

### Discussion

Consistent with Study 1, these findings revealed, using a prospective design, material and social indicators of deprivation at the individual level, and more strictly adjusted models, that 1) BMI partly mediated the association of both material deprivation and social deprivation with physical activity and 2) that this mediating pathway was moderated by gender. Specifically, the strength of the association of material and social deprivation with BMI was stronger among women, relative to men.

## General discussion

### Main findings

Findings from these two large-scale studies showed that BMI partly mediated the association between deprivation and physical activity, which extends our understanding of the pathways through which deprivation relates to physical activity. Moreover, this mediating pathway was moderated by gender, with the indirect effect of deprivation on physical activity through BMI being stronger among women, relative to men. This stronger mediating effect was explained by fact that the association between deprivation and BMI was more pronounced among women. Of note, our findings were consistent across both measures of deprivation (i.e., area deprivation in Study 1 vs perceived material and social deprivation in Study 2) and of physical activity (i.e., continuous variable in Study 1 vs dichotomous variable in Study 2), and the different designs (cross-sectional in Study 1 vs prospective in Study 3), providing robust support of evidenced associations.

### Comparison with previous studies

Our results align with past literature demonstrating that deprived adults report lower levels of physical activity in the United Kingdom (Farrell et al., 2014), as well as in Western countries (O’Donoghue et al., 2020). As hypothesized (H1), the relationship between deprivation and physical activity was partly explained by BMI, even after adjustment for age, ethnicity and occupation (Study 1) or age, country of residence, occupation, education and income (Study 2). Studies conducted among adolescents also evidenced this mediating pathway, though they assumed a reversed causal pathway. They showed that the association of deprivation and BMI was attenuated (or even non-significant), when controlling for physical activity (Clennin et al., 2020; Nevill et al., 2016). Here, we tested a reversed causal pathway (i.e., from BMI → physical activity) as recent evidence rather leads us to postulate that BMI is likely to precede physical activity. Indeed, studies testing the bi-directional association between physical activity and BMI suggested that BMI was likely to precede physical activity, than the opposite (Sagelv et al., 2021; Skrede et al., 2021). Moreover, it should be noted that the effect of physical activity on weight loss remains debated (Pontzer, 2015), especially among older adults among whom physical activity rather relates to weight maintenance than weight loss (Stephen & Janssen, 2010). While several mechanisms have been put forth to explain the detrimental effects of BMI on physical activity, spanning from physiological (e.g., higher energy expenditure, mechanical workloads) (Lafortuna et al., 2008), socio-cultural (e.g., stronger stigmatization) (Vartanian & Shaprow, 2008) to psychological pathways (e.g., more negative affective responses or lower self-efficacy) (Dikareva et al., 2016; Ekkekakis & Lind, 2006), we consider that future research is needed to disentangle the potential bi-directional relationships between BMI and physical activity. In the meantime, our findings suggest that living in a deprived area or perceiving a high level of deprivation may increase the risk of weight gain, which may in turn impede the adoption of physically active behaviors. A mechanism that may ultimately create a vicious circle, or an accumulation of disadvantages (Dannefer, 2003), on health status.

As predicted (H2), the mediating pathway between deprivation and physical activity through BMI was moderated by participants’ gender. We found consistent evidence for its moderating effect on the first component of the mediating pathway (i.e., from deprivation to BMI). In line with previous literature, we observed that, relative to men, the association of deprivation with BMI was stronger among women (Bann et al., 2017; Matheson et al., 2008). Although remaining speculative, several explanations can be put forth to explain the stronger relationship between deprivation and BMI observed among women. First, from a sociocultural lens, it has been proposed that women reporting a high level of deprivation may be less likely to experience injunctions to thinness than women living in a more privileged milieu (McLaren & Kuh, 2004; Swami et al., 2010), especially at older age (Hajek & König, 2018). In turn, this weaker cultural pressure may expose deprived women to increases in BMI across aging (von Lengerke & Mielck, 2012). Conversely, irrespectively from socio-economic circumstances, men are exposed to weaker injunctions to thinness (McLaren & Kuh, 2004; Swami et al., 2010). From a social perspective, being a women (vs a men) and experiencing a high deprivation (vs a low) increases odds of isolation (Nicolaisen & Thorsen, 2014), which could in turn foster weight gains due to the absence of social support (Kiernan et al., 2012). Finally, at the behavioral level, compared to men or to privileged women, women living in a disadvantaged environment are at higher risk to report a greater tendency to unhealthy food behaviors (e.g., skipping meal, binge eating) (Jeffery & French, 1996; Reagan & Hersch, 2005), which are related to a higher BMI (French et al., 2012). Again, these explanations would deserve to be tested by future studies, with the aim of shedding light on the mechanisms behind the exacerbated association between deprivation and BMI among women.

Importantly, across our two studies, we found evidence for a partial mediation of the association between deprivation and physical activity by BMI, with this latter explaining between 1.2% and 8.1% of this association. This suggests that other factors may conjointly underlie the association between deprivation and physical activity. While the role of environmental (e.g., access to facilities, walkability, adverse neighborhood conditions, air pollution) (Chaparro et al., 2018; Hillsdon et al., 2007; Schneider et al., 2019), cultural (e.g., class habitus) (Bourdieu, 1979; Cockerham et al., 2020; Pampel et al., 2010), or cognitive factors (e.g., motivations, cognitive functions) (Cheval et al., 2019; Hagger & Hamilton, 2020) was already identified, future research is needed to broaden our understanding of the mechanisms through which deprivation relates to physical activity.

### Strengths and limitations

Among the main strengths of this study are the consistency of results across the two independent samples. Specifically, despite differences in the characteristics of these samples (i.e., age, % of women or mean BMI, see Table 1), moderated mediating patterns were robustly evidenced among both middle-aged British adults (Study 1) and older European adults (Study 2), which strengthens the credibility and generalizability of the current findings. Other strengths of this study include the reliance on different indicators of deprivation and physical activity, the large size of samples, as well as complementary statistical approaches to test mediating patterns. However, this study has several limitations that should be acknowledged. First, despite the time-lagged assessment between deprivation, BMI and physical activity in Study 2, we cannot rule out the possibility that BMI and physical activity might be bi-directionally associated. In the same line, a higher BMI may even lead to a greater deprivation (Howe et al., 2020). Accordingly, future research remains needed before drawing any conclusion on the causality between observed variables. Second, the self-reported assessment of physical activity lacks granularity and may have led to an overestimation of physical activity levels (Lee et al., 2011). BMI was also self-reported, which may have led to inaccuracies and triggered social desirability bias (Gorber et al., 2007). Third, the present study did not allow to disentangle leisure-time and occupational physical activity, two contexts in which physical activity levels may differ depending on individuals’ level of deprivation (especially in Study 1 in which individuals were mostly working-age adults). Fourth, regarding our inclusion criteria (i.e., having complete measures on a single timepoint in Study 1 or on three timepoints in Study 2), it should be noted that a selection bias may have occurred (e.g., over-representation of participants with a good health status), thereby reducing the representativeness of our samples. Finally, other measures of participants’ adiposity status (e.g., waist circumference, fat mass) would have been interesting to consider in relation to deprivation and physical activity (Müller et al., 2012).

## Conclusions

Overall, this study showed that 1) BMI partly mediated the association between deprivation (at both the area- and the individual level) and 2) that this mediating pathway was more pronounced among women (vs men) because of a stronger effect of deprivation on BMI among them. We hope that these findings will encourage public policies and future research in considering how deprivation and gender may interact to explain health behaviors, such as physical activity.

## Data Availability

Data are available on the following websites : http://www.share-project.org/dataaccess.html and https://osf.io/uzbw9/

