## Supplementary material for "Gender Moderates the Mediating Pathway between Social Deprivation, Body Mass Index and Physical Activity"

**Table 1.** Estimates for all predictors in the models including area deprivation (Study 1).

**Table 2.** Estimates for all predictors in the models including material deprivation (Study 2).

**Table 3.** Estimates for all predictors in the models including social deprivation (Study 2).

**Table 1.** Estimates for all predictors in the models including area deprivation (Study 1).

| | Model 1a (BMI as outcome) | | | Model 1b (BMI as outcome) | | | Model 2a (PA as outcome) | | | Model 2b (PA as outcome) | | |
|---|---|---|---|---|---|---|---|---|---|---|---|---|
| **Predictors** | **b** | **95% CI** | ***p*** | **b** | **95% CI** | ***p*** | **b** | **95% CI** | ***p*** | **b** | **95% CI** | ***p*** |
| Intercept | -0.52 | -1.40 – 0.35 | 0.240 | -0.33 | -1.20 – 0.54 | 0.461 | -0.38 | -1.21 – 0.45 | 0.372 | -0.42 | -1.25 – 0.41 | 0.322 |
| Area deprivation | 0.19 | 0.16 – 0.22 | <0.001 | 0.15 | 0.12 – 0.18 | <0.001 | -0.29 | -0.32 – -0.26 | <0.001 | -0.29 | -0.32 – -0.26 | <0.001 |
| BMI | - | - | - | - | - | - | -0.05 | -0.08 – -0.03 | <0.001 | -0.05 | -0.08 – -0.02 | <0.001 |
| Gender (ref : men) | - | - | - | 0.14 | 0.08 – 0.19 | <0.001 | -0.05 | -0.08 – -0.03 | <0.001 | -0.19 | -0.24 – -0.14 | <0.001 |
| Area deprivation x gender | - | - | - | 0.17 | 0.11 – 0.22 | <0.001 | - | - | - | 0.06 | 0.00 – 0.11 | 0.035 |
| BMI x gender | - | - | - | - | - | - | | | | 0.02 | -0.04 – 0.07 | 0.505 |
| Age | -0.06 | -0.09 – -0.04 | <0.001 | -0.06 | -0.08 – -0.03 | <0.001 | -0.08 | -0.10 – -0.05 | <0.001 | -0.09 | -0.11 – -0.06 | <0.001 |
| *Employment status (ref : Carer)* | | | | | | | | | | | | |
| Employed full time | 0.69 | -0.18 – 1.56 | 0.122 | 0.47 | -0.40 – 1.34 | 0.291 | 0.37 | -0.46 – 1.20 | 0.384 | 0.43 | -0.40 – 1.26 | 0.306 |
| Employed full time / part time | 0.65 | -0.22 – 1.52 | 0.145 | 0.40 | -0.46 – 1.27 | 0.361 | 0.07 | -0.76 – 0.90 | 0.867 | 0.14 | -0.69 – 0.96 | 0.747 |
| Employed part time | 0.45 | -0.43 – 1.33 | 0.314 | 0.23 | -0.65 – 1.11 | 0.607 | 0.47 | -0.37 – 1.30 | 0.273 | 0.58 | -0.25 – 1.42 | 0.169 |
| Long-term sick/disabled | 0.89 | -0.51 – 2.29 | 0.211 | 0.72 | -0.68 – 2.11 | 0.314 | 0.26 | -1.07 – 1.59 | 0.706 | 0.19 | -1.13 – 1.52 | 0.774 |
| Looking after home / family full after home / family full | 0.25 | -0.71 – 1.21 | 0.609 | 0.02 | -0.94 – 0.98 | 0.965 | 0.24 | -0.68 – 1.15 | 0.612 | 0.38 | -0.53 – 1.30 | 0.410 |
| Other | 0.64 | -0.25 – 1.54 | 0.159 | 0.42 | -0.47 – 1.32 | 0.350 | 0.10 | -0.75 – 0.95 | 0.816 | 0.16 | -0.69 – 1.01 | 0.708 |

| | | | | | | | | | | | | |
|---|---|---|---|---|---|---|---|---|---|---|---|---|
| Permanently sick | 0.78 | -0.22 – 1.78 | 0.127 | 0.57 | -0.43 – 1.57 | 0.267 | -0.03 | -0.98 – 0.92 | 0.951 | 0.10 | -0.85 – 1.05 | 0.829 |
| Retired | 0.50 | -0.37 – 1.37 | 0.264 | 0.26 | -0.61 – 1.13 | 0.562 | 0.31 | -0.51 – 1.14 | 0.456 | 0.39 | -0.43 – 1.22 | 0.351 |
| Self-employed | 0.40 | -0.51 – 1.32 | 0.386 | 0.17 | -0.74 – 1.09 | 0.710 | 1.02 | 0.15 – 1.89 | 0.021 | 1.06 | 0.19 – 1.93 | 0.017 |
| Student | 0.42 | -0.68 – 1.51 | 0.455 | 0.17 | -0.93 – 1.26 | 0.763 | -0.29 | -1.34 – 0.75 | 0.581 | -0.30 | -1.34 – 0.74 | 0.569 |
| Unemployed | 0.59 | -0.28 – 1.46 | 0.185 | 0.39 | -0.48 – 1.26 | 0.375 | 0.10 | -0.73 – 0.93 | 0.806 | 0.15 | -0.68 – 0.97 | 0.728 |
| Unknown | 0.65 | -0.21 – 1.51 | 0.141 | 0.42 | -0.44 – 1.28 | 0.337 | 0.23 | -0.59 – 1.05 | 0.579 | 0.29 | -0.53 – 1.11 | 0.483 |
| Volunteer | 1.77 | -0.33 – 3.86 | 0.098 | 1.49 | -0.59 – 3.58 | 0.161 | -0.26 | -2.25 – 1.73 | 0.799 | -0.14 | -2.12 – 1.84 | 0.889 |
| *Ethnicity (ref : African)* | | | | | | | | | | | | |
| Arab | 0.17 | -1.18 – 1.53 | 0.801 | 0.14 | -1.21 – 1.49 | 0.837 | -0.15 | -1.44 – 1.14 | 0.815 | -0.21 | -1.49 – 1.07 | 0.751 |
| Bangladeshi | -0.33 | -1.20 – 0.53 | 0.450 | -0.19 | -1.06 – 0.67 | 0.658 | 0.48 | -0.35 – 1.30 | 0.257 | 0.41 | -0.41 – 1.22 | 0.332 |
| British | -0.02 | -0.18 – 0.15 | 0.846 | -0.01 | -0.17 – 0.16 | 0.924 | 0.46 | 0.31 – 0.62 | <0.001 | 0.47 | 0.32 – 0.63 | <0.001 |
| Caribbean | -0.01 | -0.18 – 0.16 | 0.901 | -0.04 | -0.21 – 0.13 | 0.648 | -0.00 | -0.17 – 0.16 | 0.985 | 0.02 | -0.15 – 0.18 | 0.853 |
| Chinese | -0.73 | -1.53 – 0.06 | 0.072 | -0.78 | -1.57 – 0.02 | 0.055 | 0.03 | -0.73 – 0.78 | 0.948 | 0.05 | -0.70 – 0.81 | 0.887 |
| Indian | 0.14 | -0.26 – 0.54 | 0.490 | 0.11 | -0.28 – 0.51 | 0.573 | -0.10 | -0.48 – 0.28 | 0.594 | -0.05 | -0.42 – 0.33 | 0.809 |
| Irish | -0.32 | -0.83 – 0.20 | 0.227 | -0.33 | -0.84 – 0.18 | 0.206 | 0.31 | -0.18 – 0.80 | 0.218 | 0.28 | -0.21 – 0.77 | 0.259 |
| Other Asian | -0.19 | -0.58 – 0.19 | 0.330 | -0.24 | -0.63 – 0.14 | 0.219 | 0.02 | -0.34 – 0.39 | 0.895 | 0.05 | -0.31 – 0.42 | 0.778 |
| Other Black | -0.33 | -0.65 – -0.01 | 0.045 | -0.35 | -0.67 – -0.03 | 0.031 | -0.02 | -0.32 – 0.29 | 0.908 | -0.01 | -0.31 – 0.30 | 0.963 |
| Other Ethnic | -0.78 | -1.23 – -0.33 | 0.001 | -0.86 | -1.30 – -0.41 | <0.001 | -0.02 | -0.44 – 0.41 | 0.939 | 0.00 | -0.42 – 0.43 | 0.994 |
| Other Mixed | -0.52 | -1.06 – 0.03 | 0.065 | -0.52 | -1.06 – 0.03 | 0.062 | -0.03 | -0.55 – 0.49 | 0.913 | -0.03 | -0.54 – 0.49 | 0.923 |
| Other White | -0.26 | -0.54 – 0.03 | 0.077 | -0.26 | -0.54 – 0.02 | 0.072 | 0.03 | -0.24 – 0.30 | 0.813 | 0.03 | -0.23 – 0.30 | 0.801 |
| Pakistani | 0.03 | -0.70 – 0.77 | 0.926 | -0.02 | -0.75 – 0.71 | 0.961 | -0.14 | -0.84 – 0.56 | 0.701 | -0.11 | -0.80 – 0.59 | 0.762 |
| Unknown/ Withheld | -0.12 | -0.27 – 0.02 | 0.101 | -0.13 | -0.27 – 0.02 | 0.088 | 0.11 | -0.02 – 0.25 | 0.107 | 0.12 | -0.02 – 0.25 | 0.098 |
| White and Asian | -0.84 | -1.95 – 0.27 | 0.138 | -0.87 | -1.97 – 0.24 | 0.125 | -0.61 | -1.66 – 0.45 | 0.262 | -0.60 | -1.65 – 0.46 | 0.266 |
| White and Black African | -0.00 | -0.52 – 0.51 | 0.990 | 0.06 | -0.46 – 0.57 | 0.828 | 0.26 | -0.23 – 0.75 | 0.306 | 0.23 | -0.26 – 0.71 | 0.364 |
| White and Black Caribbean | -0.19 | -0.59 – 0.20 | 0.335 | -0.20 | -0.59 – 0.19 | 0.308 | -0.06 | -0.44 – 0.31 | 0.734 | -0.06 | -0.43 – 0.31 | 0.741 |

*Note:* BMI: body mass index; PA: Physical activity status. Model 1b and Model 2b included interaction terms between gender and area deprivation on BMI (Model 1b) and between gender and area deprivation and BMI and gender on physical activity (Model 2b). Unstandardized b coefficients their 95% confidence intervals (95% CI) are reported.

**Table 2**. Estimates for all predictors in the models including material deprivation (Study 2).

| | Model 1a (BMI as outcome) | | | Model 1b (BMI as outcome) | | | Model 2a (PA as outcome) | | | Model 2b (PA as outcome) | | |
|---|---|---|---|---|---|---|---|---|---|---|---|---|
| **Predictors** | **b** | **95% CI** | ***p*** | **b** | **95% CI** | ***p*** | **OR** | **95% CI** | ***p*** | **OR** | **95% CI** | ***p*** |
| Intercept | 0.08 | -0.01 – 0.18 | 0.071 | 0.05 | -0.04 – 0.14 | 0.261 | 2.60 | 2.07 – 3.28 | <0.001 | 2.57 | 2.04 – 3.23 | <0.001 |
| Material deprivation | 0.06 | 0.02 – 0.09 | 0.001 | 0.05 | 0.02 – 0.08 | 0.004 | 0.86 | 0.79 – 0.92 | <0.001 | 0.86 | 0.80 – 0.93 | <0.001 |
| BMI | | | | | | | 0.82 | 0.78 – 0.87 | <0.001 | 0.81 | 0.76 – 0.86 | <0.001 |
| Gender (ref : men) | | | | -0.17 | -0.22 – -0.13 | <0.001 | | | | 0.80 | 0.72 – 0.89 | <0.001 |
| Material deprivation x gender | | | | 0.08 | 0.03 – 0.13 | 0.001 | | | | 0.95 | 0.85 – 1.07 | 0.391 |
| BMI x gender | | | | | | | | | | 1.07 | 0.96 – 1.20 | 0.220 |
| Age | -0.11 | -0.13 – -0.08 | <0.001 | -0.12 | -0.14 – -0.09 | <0.001 | 0.53 | 0.49 – 0.57 | <0.001 | 0.52 | 0.49 – 0.56 | <0.001 |
| *Country (ref: Belgium)* | | | | | | | | | | | | |
| Austria | 0.16 | 0.05 – 0.27 | 0.004 | 0.18 | 0.07 – 0.29 | 0.002 | 1.34 | 1.03 – 1.76 | 0.031 | 1.36 | 1.04 – 1.79 | 0.025 |
| Czech Republic | 0.19 | 0.10 – 0.28 | <0.001 | 0.20 | 0.11 – 0.29 | <0.001 | 1.46 | 1.18 – 1.82 | 0.001 | 1.49 | 1.20 – 1.86 | <0.001 |
| Denmark | -0.13 | -0.21 – -0.06 | 0.001 | -0.13 | -0.20 – -0.05 | 0.001 | 1.86 | 1.51 – 2.30 | <0.001 | 1.88 | 1.52 – 2.32 | <0.001 |
| France | -0.14 | -0.22 – -0.05 | 0.001 | -0.13 | -0.21 – -0.05 | 0.002 | 1.64 | 1.33 – 2.03 | <0.001 | 1.66 | 1.34 – 2.05 | <0.001 |
| Germany | 0.11 | 0.03 – 0.20 | 0.011 | 0.11 | 0.03 – 0.20 | 0.009 | 1.61 | 1.29 – 2.01 | <0.001 | 1.61 | 1.30 – 2.02 | <0.001 |
| Italy | -0.17 | -0.25 – -0.09 | <0.001 | -0.18 | -0.26 – -0.10 | <0.001 | 0.83 | 0.69 – 0.99 | 0.041 | 0.81 | 0.67 – 0.97 | 0.024 |
| Spain | 0.11 | 0.02 – 0.20 | 0.014 | 0.10 | 0.01 – 0.19 | 0.023 | 1.51 | 1.22 – 1.87 | <0.001 | 1.48 | 1.20 – 1.83 | <0.001 |
| Sweden | -0.06 | -0.15 – 0.02 | 0.129 | -0.05 | -0.13 – 0.04 | 0.281 | 2.35 | 1.88 – 2.94 | <0.001 | 2.40 | 1.92 – 3.01 | <0.001 |
| Switzerland | -0.26 | -0.35 – -0.17 | <0.001 | -0.25 | -0.34 – -0.15 | <0.001 | 1.76 | 1.38 – 2.25 | <0.001 | 1.78 | 1.40 – 2.28 | <0.001 |
| *Income (ref: Easily)* | | | | | | | | | | | | |
| Fairly easily | 0.10 | 0.05 – 0.15 | <0.001 | 0.11 | 0.06 – 0.16 | <0.001 | 0.79 | 0.70 – 0.90 | <0.001 | 0.79 | 0.70 – 0.90 | <0.001 |
| With some difficulty | 0.14 | 0.07 – 0.21 | <0.001 | 0.15 | 0.08 – 0.22 | <0.001 | 0.71 | 0.60 – 0.84 | <0.001 | 0.71 | 0.60 – 0.84 | <0.001 |
| With great difficulty | 0.25 | 0.13 – 0.37 | <0.001 | 0.25 | 0.14 – 0.37 | <0.001 | 0.74 | 0.57 – 0.98 | 0.035 | 0.75 | 0.57 – 0.99 | 0.042 |

| | | | | | | | | | | | | |
|---|---|---|---|---|---|---|---|---|---|---|---|---|
| *Education (ref: Primary)* | | | | | | | | | | | | |
| Secondary | -0.10 | -0.16 – -0.04 | 0.001 | -0.10 | -0.16 – -0.05 | <0.001 | 1.07 | 0.93 – 1.22 | 0.349 | 1.06 | 0.92 – 1.21 | 0.408 |
| Tertiary | -0.27 | -0.35 – -0.20 | <0.001 | -0.27 | -0.34 – -0.20 | <0.001 | 1.35 | 1.12 – 1.63 | 0.002 | 1.35 | 1.11 – 1.63 | 0.002 |
| *Occupation status (ref: High skill)* | | | | | | | | | | | | |
| Low skill | 0.00 | -0.05 – 0.06 | 0.909 | 0.05 | -0.00 – 0.10 | 0.072 | 0.97 | 0.84 – 1.11 | 0.630 | 1.01 | 0.87 – 1.16 | 0.927 |
| Never worked | -0.07 | -0.17 – 0.04 | 0.199 | 0.04 | -0.07 – 0.15 | 0.452 | 0.65 | 0.51 – 0.82 | <0.001 | 0.74 | 0.58 – 0.96 | 0.021 |

*Note:* BMI: body mass index; PA: Physical activity status. Model 1b and Model 2b included interaction terms between gender and material deprivation on BMI (Model 1b) and between gender and material deprivation and BMI and gender on physical activity (Model 2b). Unstandardized b coefficients and odds-ratios (OR) and their 95% confidence intervals (95% CI) are reported.

**Table 3.** Estimates for all predictors in the models including social deprivation (Study 2).

| | Model 1a (BMI as outcome) | | | Model 1b (BMI as outcome) | | | Model 2a (PA as outcome) | | | Model 2b (PA as outcome) | | |
|---|---|---|---|---|---|---|---|---|---|---|---|---|
| **Predictors** | **b** | **95% CI** | ***p*** | **b** | **95% CI** | ***p*** | **OR** | **95% CI** | ***p*** | **OR** | **95% CI** | ***p*** |
| Intercept | 0.06 | -0.03 – 0.15 | 0.192 | 0.03 | -0.06 – 0.12 | 0.498 | 2.69 | 2.15 – 3.38 | <0.001 | 2.66 | 2.12 – 3.35 | <0.001 |
| Social deprivation | 0.04 | 0.02 – 0.06 | 0.001 | 0.04 | 0.02 – 0.06 | 0.001 | 0.74 | 0.70 – 0.79 | <0.001 | 0.75 | 0.71 – 0.80 | <0.001 |
| BMI | - | - | - | - | - | - | 0.83 | 0.78 – 0.87 | <0.001 | 0.81 | 0.77 – 0.86 | <0.001 |
| Gender (ref : men) | - | - | - | -0.19 | -0.23 – -0.15 | <0.001 | - | - | - | 0.83 | 0.75 – 0.93 | 0.001 |
| Social deprivation x gender | - | - | - | 0.05 | 0.01 – 0.10 | 0.015 | - | - | - | 0.95 | 0.85 – 1.05 | 0.319 |
| BMI x gender | - | - | - | | | | - | - | - | 1.07 | 0.96 – 1.19 | 0.233 |
| Age | -0.11 | -0.14 – -0.09 | <0.001 | -0.12 | -0.15 – -0.10 | <0.001 | 0.55 | 0.52 – 0.59 | <0.001 | 0.55 | 0.51 – 0.59 | <0.001 |
| *Country (ref: Belgium)* | | | | | | | | | | | | |
| Austria | 0.18 | 0.07 – 0.29 | 0.001 | 0.19 | 0.08 – 0.30 | 0.001 | 1.21 | 0.93 – 1.59 | 0.163 | 1.23 | 0.94 – 1.62 | 0.137 |
| Czech Republic | 0.18 | 0.09 – 0.27 | <0.001 | 0.20 | 0.11 – 0.29 | <0.001 | 1.63 | 1.31 – 2.04 | <0.001 | 1.65 | 1.32 – 2.06 | <0.001 |
| Denmark | -0.12 | -0.19 – -0.04 | 0.003 | -0.11 | -0.19 – -0.03 | 0.006 | 1.67 | 1.35 – 2.06 | <0.001 | 1.68 | 1.36 – 2.08 | <0.001 |
| France | -0.13 | -0.21 – -0.05 | 0.002 | -0.12 | -0.21 – -0.04 | 0.003 | 1.64 | 1.33 – 2.03 | <0.001 | 1.66 | 1.34 – 2.05 | <0.001 |
| Germany | 0.13 | 0.04 – 0.21 | 0.004 | 0.13 | 0.04 – 0.21 | 0.003 | 1.54 | 1.24 – 1.93 | <0.001 | 1.55 | 1.24 – 1.93 | <0.001 |
| Italy | -0.17 | -0.25 – -0.09 | <0.001 | -0.19 | -0.26 – -0.11 | <0.001 | 0.87 | 0.73 – 1.05 | 0.156 | 0.86 | 0.71 – 1.04 | 0.114 |
| Spain | 0.12 | 0.04 – 0.21 | 0.005 | 0.11 | 0.02 – 0.20 | 0.013 | 1.47 | 1.19 – 1.82 | <0.001 | 1.45 | 1.18 – 1.79 | 0.001 |
| Sweden | -0.05 | -0.14 – 0.03 | 0.211 | -0.03 | -0.12 – 0.05 | 0.428 | 2.22 | 1.77 – 2.79 | <0.001 | 2.26 | 1.81 – 2.84 | <0.001 |
| Switzerland | -0.25 | -0.34 – -0.15 | <0.001 | -0.23 | -0.32 – -0.14 | <0.001 | 1.64 | 1.29 – 2.10 | <0.001 | 1.66 | 1.30 – 2.13 | <0.001 |
| *Income (ref: Easily)* | | | | | | | | | | | | |
| Fairly easily | 0.11 | 0.06 – 0.16 | <0.001 | 0.11 | 0.06 – 0.16 | <0.001 | 0.80 | 0.70 – 0.91 | 0.001 | 0.80 | 0.70 – 0.91 | 0.001 |
| With some difficulty | 0.18 | 0.11 – 0.24 | <0.001 | 0.19 | 0.12 – 0.25 | <0.001 | 0.67 | 0.58 – 0.79 | <0.001 | 0.68 | 0.58 – 0.79 | <0.001 |
| With great difficulty | 0.33 | 0.23 – 0.43 | <0.001 | 0.33 | 0.23 – 0.43 | <0.001 | 0.69 | 0.55 – 0.87 | 0.002 | 0.69 | 0.55 – 0.87 | 0.002 |

| | | | | | | | | | | | | |
|---|---|---|---|---|---|---|---|---|---|---|---|---|
| *Education (ref: Primary)* | | | | | | | | | | | | |
| Secondary | -0.10 | -0.15 – -0.04 | 0.001 | -0.10 | -0.16 – -0.05 | <0.001 | 1.03 | 0.90 – 1.18 | 0.644 | 1.03 | 0.90 – 1.18 | 0.708 |
| Tertiary | -0.27 | -0.34 – -0.20 | <0.001 | -0.26 | -0.34 – -0.19 | <0.001 | 1.27 | 1.05 – 1.54 | 0.013 | 1.27 | 1.05 – 1.54 | 0.013 |
| *Occupation status (ref: High skill)* | | | | | | | | | | | | |
| Low skill | 0.00 | -0.05 – 0.05 | 0.971 | 0.04 | -0.01 – 0.10 | 0.099 | 1.00 | 0.87 – 1.14 | 0.958 | 1.03 | 0.89 – 1.19 | 0.669 |
| Never worked | -0.07 | -0.18 – 0.03 | 0.155 | 0.04 | -0.07 – 0.15 | 0.465 | 0.70 | 0.55 – 0.90 | 0.004 | 0.79 | 0.62 – 1.02 | 0.074 |

*Note:* Model 1b and Model 2b included an interaction terms between gender and social deprivation on BMI (Model 1b), between gender and social deprivation and BMI and gender on physical activity (Model 2b). Unstandardized b coefficients and odds-ratios (OR) and their 95% confidence intervals (95% CI) are reported.
